# Survival analysis of coronary care unit patients from MIMIC IV database

**DOI:** 10.1101/2022.04.29.22274467

**Authors:** Pedro G. Lanzieri, Dayanna Q. Palmer, Ronaldo A. Gismondi, Valéria T. Baltar, Flavio L. Seixas

## Abstract

The profile of hospitalizations in coronary care units (CCU) includes patients with different age groups, multiple comorbidities and causes of hospitalization that may or may not be primarily cardiac. This study aimed to estimate survival time and evaluate the association and impact of different factors on this time in a cohort of patients admitted to CCU. A cohort of 7120 adult patients admitted to CCU was analyzed from a subset of data from the MIMIC-IV database (Medical Information Mart for Intensive Care, version 4). A descriptive analysis was performed using Kaplan-Meier survival analysis, with a Log-rank test to establish comparisons between groups. Survival regression was modeled using Cox’s proportional risk models for the multiple analysis. The p-value was defined as ¡ 0.05 as statistically significant. In patients who died during hospitalization, there was a higher average age, longer hospital stay, and a higher rate of heart and respiratory rate, all with p ¡ 0.001. Median overall survival was 28 days (95% CI 26-30 days). The survival probability curve presented a higher inclination in the first weeks, reaching a stable value close to 20% at 10 weeks after hospitalization. When Cox’s regression adjusted for age, gender and comorbidities was performed, hyperpotassemia was shown to be an independent risk factor for in-hospital mortality (RR = 1.22, 95% CI: 1.14-1.30) in this group of patients. These results reinforced that the electronic health record may contain, already in the first hours of hospitalization, relevant information to understand the progression of diseases and identify future directions for research. This study is expected to clarify important topics related to the MIMIC-IV database and enable further research using this patient database. Knowledge of the characteristics of the CCU population can allow better management of physical and human hospital resources.

## Introduction

Cardiovascular diseases are among the leading causes of death in the world. To improve outcomes in these cases, specific intensive care is required. [1] The profile of hospitalizations in the Coronary Care Units (CCU) includes patients with different age groups, multiple comorbidities, and causes of hospitalization that may or may not be primarily cardiac. [2]

The diverse characteristics of these patients make their classification into risk stratification groups challenging. This occurs because of the difficulty of generalizing data collected from other contexts, such as the CCUs, and also because of the need to reevaluate the applicability of markers previously studied in other contexts (for example, non-cardiac intensive care patients). [3] CCUs are of even greater research interest because, to date, risk prediction tools designed specifically for this population and prospectively validated in different cardiac units are not yet available for clinical practice [4], and medical societies have encouraged research to guide evidence-based conduct. [5]

Professionals working in intensive care units often need to make decisions quickly, based on their clinical judgment. For this reason, all knowledge capable of helping the understanding of factors that increase or decrease the criticality of a clinical condition can help them in decision making. [6] This includes the use of electronic health record (EHR) data to characterize the patient’s health status, also to predict future outcomes, including length of hospital stay and in-hospital mortality. [7]

Several clinical data and complementary tests have already been studied as mortality risk factors among critically ill patients, and this knowledge is used for better management of physical and human hospital resources. [8] However, differences between populations can alter the effect of certain factors, so that the most reliable - although difficult in practical terms - is to define the specific risk for each population and in each context. [9] In addition, the evaluation of medical record data may allow the identification of high-risk individuals. Survival analysis may be useful in these cases to analyze the time to the event of interest (death) in order to estimate the expected survival time, given the clinical observations of the patient. [10]

On the other hand, little progress has been made in models that apply this knowledge to a great proportion of hospitals due to several challenges, which stand out: the lack of registration data in EHR format, and the lack of interoperability of data and medical record systems. Added to this is the technical challenge of understanding the concepts involved in the survival analysis and also of dealing with unbalanced data, as often occurs in factors such as hospital mortality and length of hospital stay. [11]

This study aimed to estimate the expected survival time, and to evaluate the association and impact of different factors for the survival time of hospitalized patients, based on a cohort of patients hospitalized in CCUs.

The rest of this article is organized as follows: in Section 2, the methodology of the research is described; in Section 3, the results obtained are presented; in Section 4 the discussion is made and in Section 5 the study is concluded.

## Materials and methods

### Study Population

Retrospective, unicentric study of a cohort of patients aged 18 years or older admitted to the Beth Israel Deaconess Medical Center, Boston, MA, USA, between 2008 and 2019.

The inclusion criteria were:

1. Age 18 years old or older;
2. Admissions to CCU.

The criteria of exclusion were:

1. Records with invalid or incomplete information for that research, i.e., no heart rate measurements, or incomplete administrative procedures, no ICU admission or discharge record;
2. Organ donors, which are often registered as “readmissions” for administrative purposes;
3. Hospitalizations lasting less than 4 hours, because they are usually preoperative or for subsequent transfer to another section of the hospital.

This study was conducted under the waiver of informed consent for presenting minimal risk to patients. The identification data has been hidden for privacy guarantees, and the investigation is in accordance with the principles described in the Helsinki Declaration.

After the completion of the National Institutes of Health online training and the Protecting Human Research Participants exame (no. 45395590), Permission has been granted to access MIMIC-IV data.

The MIMIC project was approved by Beth Israel Deaconess Medical Center and by the Massachusetts Institute of Technology Institutional Review Board (Cambridge, Massachusetts).

### Description of clinical data

The diagnoses were determined using the International Classification of Diseases, Ninth Revision (ICD-9). For cardiac diagnoses, the following codes were considered: Myocardial Infarction: I21, I22, I252; Heart failure: I099, I110, I130, I132, I255, I420, I425, I426, I427, I428, I429, P290; Peripheral Artery Disease: I70, I71, I731, I738, I739, I771, I790, I792, K551, K558, K559, Z958, Z959; and Cerebrovascular Disease: G45, G46, I60 to I69, H340.

Hospitalization in CCU comprised patients who were admitted and discharged from the coronary care unit, regardless of the admission diagnosis. The concept of length of stay was characterized by the time difference, in days, between admission and hospital discharge.

In-hospital mortality was defined by the death record in the medical record at a date included in the period of hospitalization.

The clinical data analyzed are presented in Table 1. The vital signs of admission were characterized as the first recorded value after, or closer to, the moment of admission to the CCU. Laboratory tests were evaluated according to the reference values and units of measurement provided by the EHR, with two decimal places of accuracy (when necessary). Hypocalcemia was defined as serum potassium *<* 3.5 meq/L and Hyperpotassemia ≥ 5 meq/L.

**Table 1.** Epidemiological, clinical and laboratory profile of the study population. HR: heart rate; RR: respiratory rate; SBP: systolic blood pressure; DBP: diastolic blood pressure; MBP: mean blood pressure; SO2: pulse oximetry; TAP: prothrombin time; PTT: thromboplastin time; NT-ProBNP: N-fragment terminal B-type natriuretic peptide.

|  | Absent | Total | Survivors | Deaths | p-value | Test | DP |
| --- | --- | --- | --- | --- | --- | --- | --- |
| n |  | 8035 | 6468 | 1567 |  |  |  |
| Age, median [Q1,Q3] | 0 | 72.1 [60.9,82.4] | 71.0 [59.9,81.7] | 76.6 [66.0,84.9] | 0.001 | Kruskal-Wallis | 0.340 |
| Male, n (%) | 0 | 4600 (57.2) | 3722 (57.5) | 878 (56.0) | 0.290 | Chi-squared | 0.031 |
| CCU stay length, median [Q1,Q3] | 0 | 1.9 [1.1,3.4] | 1.9 [1.0,3.1] | 2.2 [1.1,4.6] | 0.001 | Kruskal-Wallis | 0.284 |
| In-hospital stay length, median [Q1,Q3] | 0 | 6.0 [3.2,10.8] | 5.9 [3.3,10.3] | 6.6 [3.0,12.4] | 0.290 | Kruskal-Wallis | 0.082 |
| HR | 8 | 99.4 (23.1) | 98.1 (22.5) | 104.7 (24.4) | 0.001 | Two-Sample t-Test | 0.282 |
| RR | 14 | 28.0 (6.2) | 27.6 (5.9) | 29.7 (6.7) | 0.001 | Two-Sample t-Test | 0.323 |
| SBP | 63 | 143.4 (24.3) | 144.3 (23.5) | 139.5 (27.1) | 0.001 | Two-Sample t-Test | -0.190 |
| DBP | 63 | 87.4 (18.7) | 88.0 (18.2) | 84.9 (20.5) | 0.001 | Two-Sample t-Test | -0.162 |
| MBP | 14 | 102.7 (24.1) | 103.1 (22.7) | 101.1 (29.0) | 0.010 | Two-Sample t-Test | -0.078 |
| SO2 | 32 | 99.2 (1.6) | 99.2 (1.2) | 99.1 (2.7) | 0.014 | Two-Sample t-Test | -0.083 |
| Temperature | 212 | 37.1 (0.7) | 37.1 (0.6) | 37.1 (1.0) | 0.004 | Two-Sample t-Test | -0.097 |
| Glucose | 244 | 312.2 (11327.6) | 335.2 (12601.9) | 215.4 (118.5) | 0.451 | Two-Sample t-Test | -0.013 |
| Creatinine | 47 | 1.8 (1.6) | 1.6 (1.5) | 2.4 (1.9) | 0.001 | Two-Sample t-Test | 0.447 |
| Hematocrit | 60 | 35.6 (6.3) | 35.9 (6.3) | 34.3 (6.5) | 0.001 | Two-Sample t-Test | -0.245 |
| White blood count | 72 | 12.1 (7.0) | 11.6 (6.4) | 14.3 (9.0) | 0.001 | Two-Sample t-Test | 0.344 |
| Platelets | 68 | 236.3 (101.6) | 236.6 (99.1) | 234.9 (111.6) | 0.567 | Two-Sample t-Test | -0.017 |
| Sodium | 51 | 139.2 (4.6) | 139.2 (4.2) | 138.8 (5.9) | 0.008 | Two-Sample t-Test | -0.082 |
| Potassium | 50 | 4.6 (0.9) | 4.6 (0.8) | 4.9 (1.0) | 0.001 | Two-Sample t-Test | 0.330 |
| TAP | 721 | 18.2 (12.2) | 17.4 (10.9) | 21.9 (15.8) | 0.001 | Two-Sample t-Test | 0.336 |
| PTT | 763 | 59.3 (41.6) | 57.3 (40.5) | 67.3 (44.8) | 0.001 | Two-Sample t-Test | 0.233 |
| Troponin T | 3880 | 1.5 (2.9) | 1.4 (2.7) | 1.7 (3.4) | 0.031 | Two-Sample t-Test | 0.083 |
| NT-proBNP | 6541 | 8775.8 (11823.4) | 7726.5 (10917.6) | 13113.9 (14219.7) | 0.001 | Two-Sample t-Test | 0.425 |

The data set of this study is a subset of MIMIC-IV. [12] MIMIC (Medical Information Mart for Intensive Care) is a freely available database comprising unidentified data relating to the health of more than 70.000 ICU admissions.

In order to minimize the risk of associated treatment biases and readmissions in the CCU, only data from the first admission of each patient were analyzed.

The dataset required basic transformations for use in this study. At that stage, columns with missing data in proportion above 10% were removed for dimensionality reduction, in addition to the search for outliers (identified as above 3 standard deviations of the mean). In addition, additional columns were created by the one hot encoding technique (representation of the qualitative variables in binary form), to adapt the data to models that accept only numbers, rounded to two decimal places for quantitative variables.

New categories were created for grouping nominal information: (1) age, according to the following scale: 18 to 24 years old, 25 to 49 years old, 50 to 64 years old, 65 to 80 years old and above 80 years old; and (2) comorbidities of interest.

At the end of those steps, the records in which there was any missing value were filled out by the principal component analysis method, which takes into account the similarities between the observations and the relationship between the variables.

### Statistical Analysis

The event of interest taken into consideration was in-hospital death by all causes, and survival time was the duration until the event of interest occurred, that is, the length of hospital stay. A descriptive analysis was performed using the Kaplan-Meier method (KM), with a Log-rank test to establish comparisons between groups. The description of the sample of patients studied was made with frequency measurements for the qualitative variables and average and standard deviation for the continuous variables. The groups were compared through the T test of two samples for continuous variables and Chi-square test for qualitative variables. The correlation by Pearson’s method was used to evaluate collinearity, using values of 0.3 for low and 0.8 for high correlation. The value of a vital signal was considered an outlier and defined as absent if it is beyond the plausible physiological intervals based on clinical knowledge. Survival regression was modeled using the Cox proportional risk models to determine the possible association of predictive variables and to obtain the adjusted Hazard ratio. P-value ¡ 0.05 was considered significant. Due to the nature of the data set, different configurations for survival analysis and survival models (available in a collaborative notebook) have been tried out and only the most relevant results will be discussed.

Statistical analysis was performed in Python using auxiliary libraries (*lifelines* [13], *tableone* [14]).

The authors had access to the data set of this study and were responsible for data integrity and statistical analysis.

## Results

### Studied Population

Considering the 76540 admissions registered in ICU, 9541 of them were in CCU, and of these, 626 readmissions were excluded. 1795 records were treated due to the absence of one or more of the variables of interest. A total of 7120 patients were included in the study, with an average age of 70.4 years old and a proportion of 57.1% of males. The in-hospital death rate was 17.5%.

In patients who died during hospitalization, there was a higher average age, longer hospital stay, and higher heart and respiratory rate records, all with *p <* 0.001. There was no statistically significant difference for mean blood pressure, oxygen saturation or body temperature levels.

Table 1 presents the main characteristics of the population, including the groups that had or did not die in hospital.

No high correlation was found between any of the variables selected in the study. As expected, the highest indices were among the attributes related to blood pressure measurement:0.48 for both systolic blood pressure (SBP) and mean blood pressure (MBP), 0.59 between diastolic blood pressure (DBP) and MBP, and 0.48 between SBP and DBP. The correlation between length of hospital stay and length of hospital stay in CCU was 0.42. Regarding comorbidities, the correlation between kidney disease and diabetes mellitus with chronic complications obtained the highest rate, 0.37. The correlation between heart failure and myocardial infarction with mortality was only 0.07 in these patients.

The average length of stay in the CCU was 2.9 days (standard deviation: 0.2). When stratified by discharge and death, averages of 2.5 and 3.6 days of hospitalization in this sector were observed, respectively. As expected, there was a higher proportion of records on the left in the hospital admission time charts, as shown in Fig 1.

**Fig 1.**
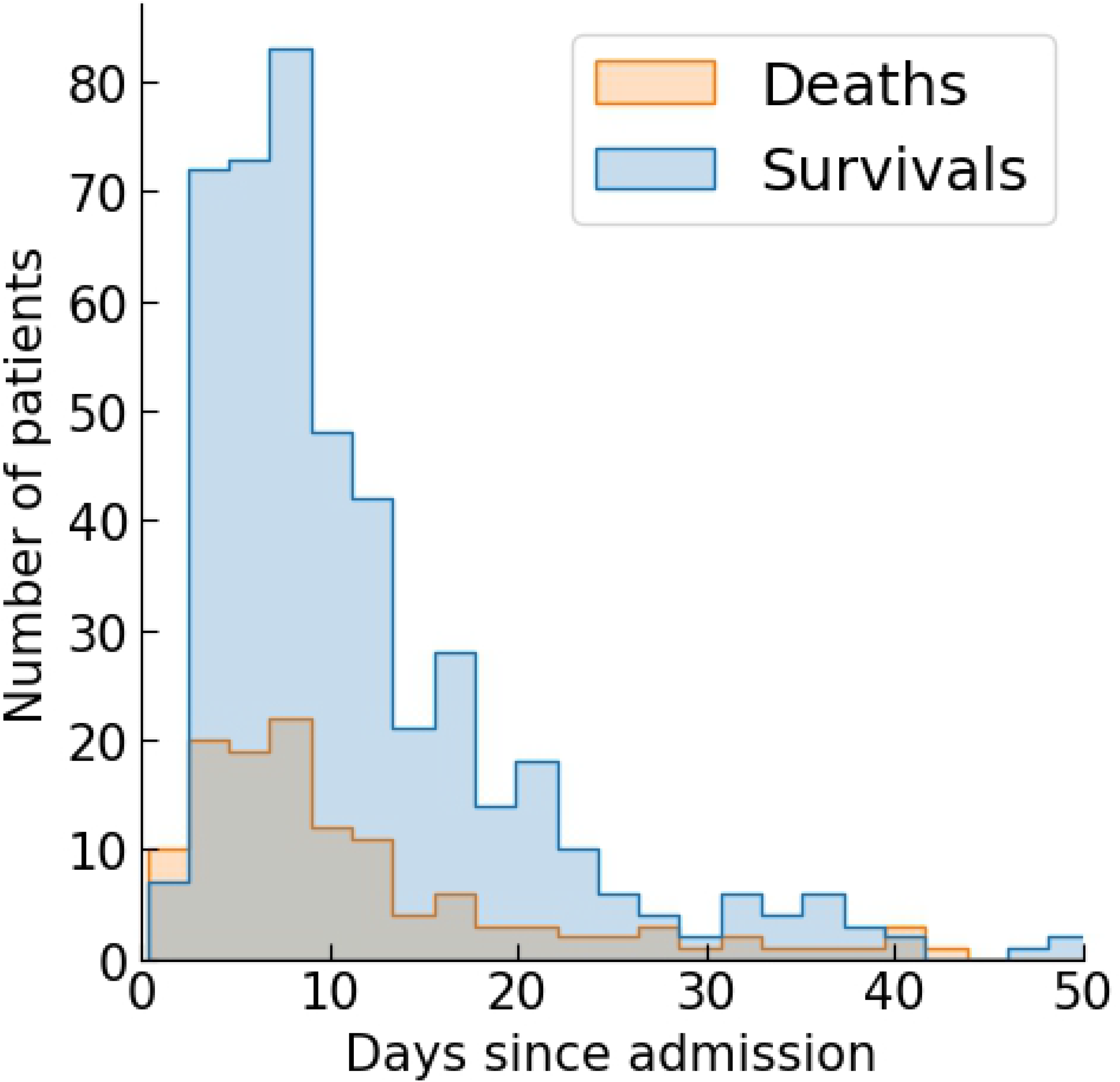
Distribution of the number of patients per day of total length of hospitalization conditioned by the occurrence or not of in-hospital death.

### Survival Analysis

At the end of the follow-up period, the overall survival estimated by the KM without stratification was 86% after 7 days, 70% after 14 days and 50% with 28 days of hospitalization. The overall survival curve is shown in Fig 2, and suggests that, after an initial period of decline, the probability of survival tends to reach a stable value around 75 days.

**Fig 2.**
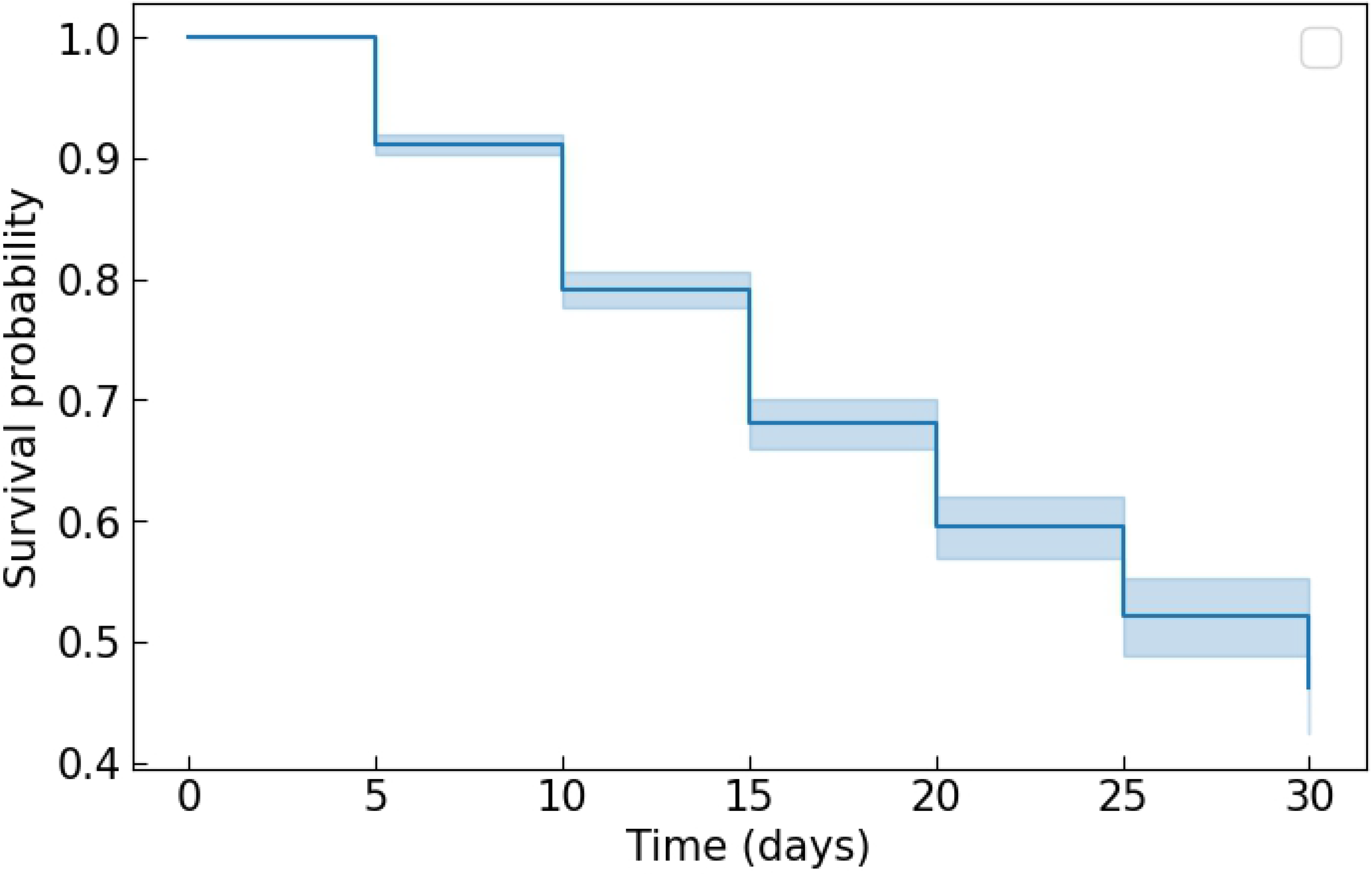
Kaplan-Meier survival curves grouped by comorbidities of interest. The shaded area represents 95% CI. In the first weeks of hospitalization, patients with these diagnoses are less likely to survive. HF: Heart failure; AMI: Acute myocardial infarction; PAD: Peripheral arterial disease; CVD: Cerebrovascular disease.

Average overall survival was 28 days (95% CI: 26-30 days). When considering gender, the values were 27 (95% CI: 25-31) for men and 26 (95% CI: 22-31) for women. Comparison with Log-rank test obtained statistical significance. Individuals who did not die during hospitalization were labeled censored on the right. When segmented according to the presence of comorbidities of interest, the probability of survival has tended to decrease in the groups with the disease, in all scenarios evaluated (Fig 3). The average survival times by comorbidities of interest are shown in Table 2.

**Table 2.** Mean survival time of different population groups based on comorbidities. The same patient may belong to more than one group. *: p-value *<* 0.05.

| Group | Length (days) | 95% CI |
| --- | --- | --- |
| **All patients (n = 7120)** | 28 | 26-30* |
| **Heart failure** |  |  |
| Present (n = 3912) | 27 | 25-31* |
| Absent | 25 | 20-30* |
| **Myocardial Infarction** |  |  |
| Present (n = 1741) | 27 | 23-29* |
| Absent | 27 | 24-32* |
| **Peripheral Arterial Disease** |  |  |
| Present (n = 710) | 25 | 21-28 |
| Absent | 28 | 25-31 |
| **Cerebrovascular Disease** |  |  |
| Present (n = 340) | 24 | 21-29* |
| Absent | 27 | 25-32* |

**Fig 3.**
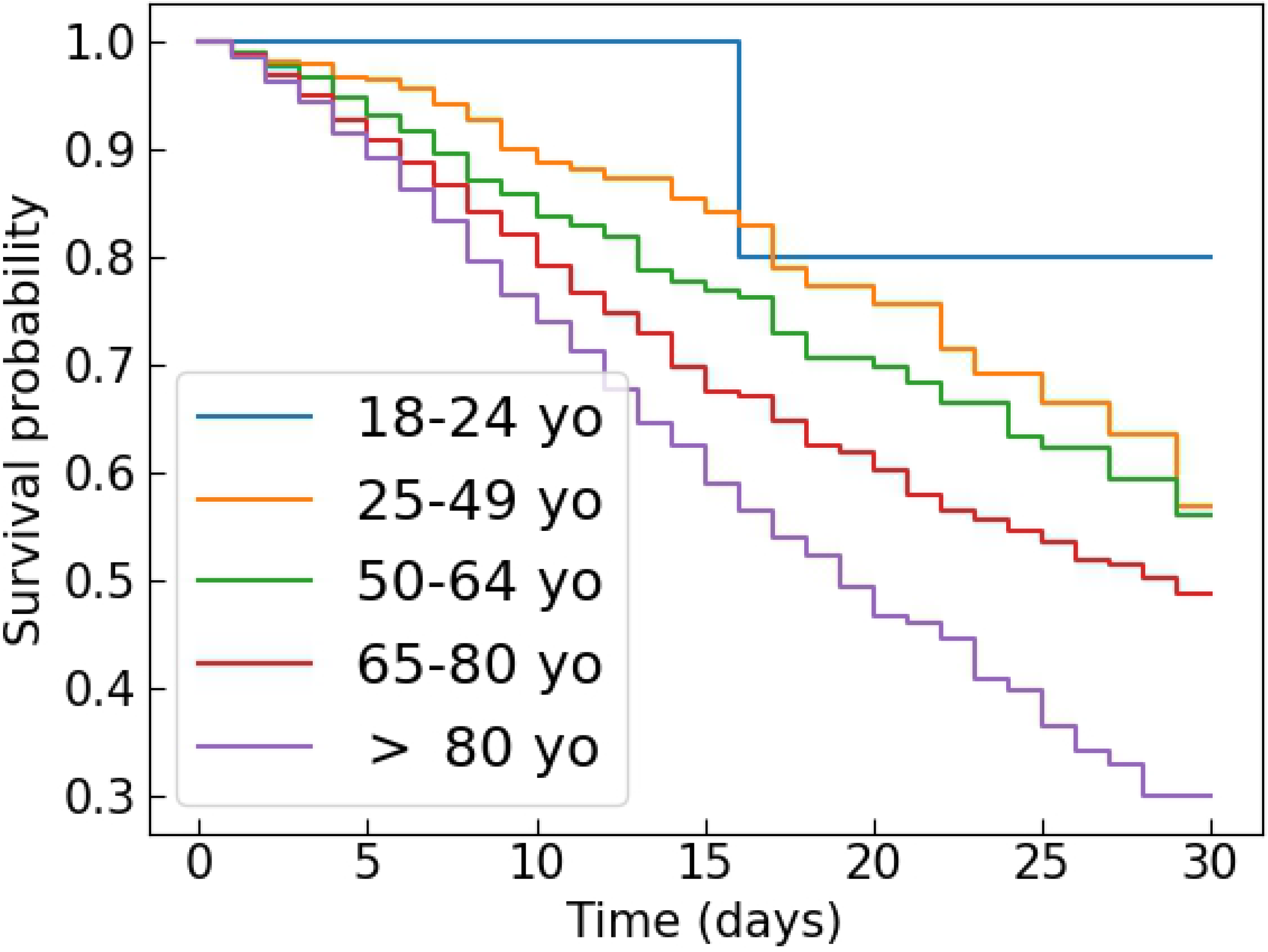
Kaplan-Meier survival curves grouped by comorbidities of interest. The shaded area represents 95% CI. In the first weeks of hospitalization, patients with these diagnoses are less likely to survive. HF: Heart failure; AMI: Acute myocardial infarction; PAD: Peripheral arterial disease; CVD: Cerebrovascular disease.

### Proportional risks analysis

When the analysis of proportional risks was performed, patients who had lower risk of death in the CCU were those who had the following characteristics: lower age, lower respiratory and heart rates (but within the values considered normal) and absence of a history of myocardial infarction, diabetes mellitus or metastasis neoplasia.

Table 3 represents the main coefficients obtained by the Cox model. The relative risk values (RR) are shown for analysis of proportional risks, with the lower and upper limits of the 95% CI. The interpretation of the results indicates that an increase of one unit in the value of the variable age will cause the baseline risk to increase by a factor of 1.02, which represents a 2% increase. In the Cox proportional risk model, a higher risk means a higher risk of occurrence of the event.

**Table 3.**
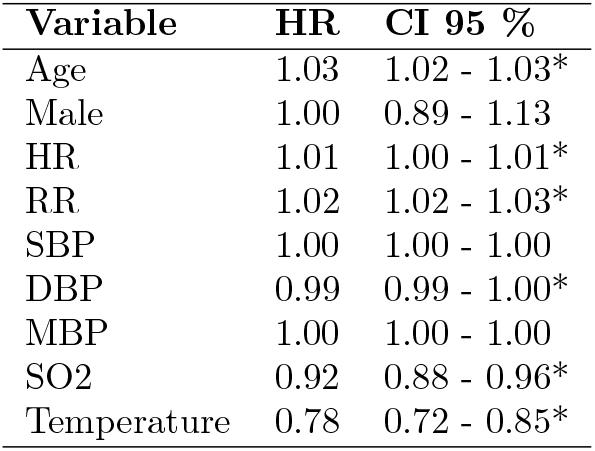
Proportional risks by the Cox regression model for the initial set of variables. *: p-value ¡ 0.05; HR: heart rate; RR: respiratory rate; SBP: systolic blood pressure; DBP: diastolic blood pressure; MAP: mean blood pressure; SO2: pulse oximetry.

| Variable | HR | CI 95 % |
| --- | --- | --- |
| Age | 1.03 | 1.02 - 1.03* |
| Male | 1.00 | 0.89 - 1.13 |
| HR | 1.01 | 1.00 - 1.01* |
| RR | 1.02 | 1.02 - 1.03* |
| SBP | 1.00 | 1.00 - 1.00 |
| DBP | 0.99 | 0.99 - 1.00* |
| MBP | 1.00 | 1.00 - 1.00 |
| SO2 | 0.92 | 0.88 - 0.96* |
| Temperature | 0.78 | 0.72 - 0.85* |

The following variables were inversely associated with in-hospital mortality by the Cox model: length of stay, diagnosis of heart failure, and higher levels of body temperature, oxygen saturation, and DBP. In these cases, the increment in the value of the quantitative variables reduces the baseline risk.

A step regression was performed to find other variables of interest that were widely available in this subgroup of patients, and, at the same time, that had physiological and statistical relevance.

586 patients were selected from the initial cohort whose medical records included laboratory variables commonly used in clinical practice. This subset of data was processed by the Cox model with adjustment for age, gender, comorbidities and vital signs.

The results are shown in Table 4. The most significant risk was increased serum potassium levels as an independent risk factor for in-hospital mortality (RR = 1.22, 95% CI: 1.14-1.30). Creatinine, hematocrit and leukocytosis were also found to be representative variables of risk of death, but with less contribution to the model.

**Table 4.** Proportional risks, with additional variables, according to the Cox model adjusted for age, gender and comorbidities. *: p-value *<* 0.05.

| Variable | HR | 95% CI |
| --- | --- | --- |
| Glucose | 1.00 | 1.00-1.00 |
| Creatinine | 1.07 | 1.03-1.10* |
| Hematocrit | 1.07 | 1.03-1.11* |
| White blood cells count | 1.02 | 1.01-1.02* |
| Platelets | 1.00 | 1.00-1.00 |
| Sodium | 1.00 | 0.99-1.01 |
| Potassium | 1.22 | 1.14-1.30* |
| TAP | 1.00 | 0.99-1.02 |
| PTT | 1.00 | 1.00-1.00 |
| Troponin T | 1.04 | 0.96-1.13 |
| NT-ProBNP | 1.00 | 1.00-1.00 |

These findings allowed a graphical representation of the curves as the value of when a single variable was changed, which depicts the impact of the variation of potassium levels in different patient admission scenarios. Fig 4 depicts the proportional risks represented by potassium disorders, in particular the increased risk in patients with severe hyperpotassemia.

**Fig 4.**
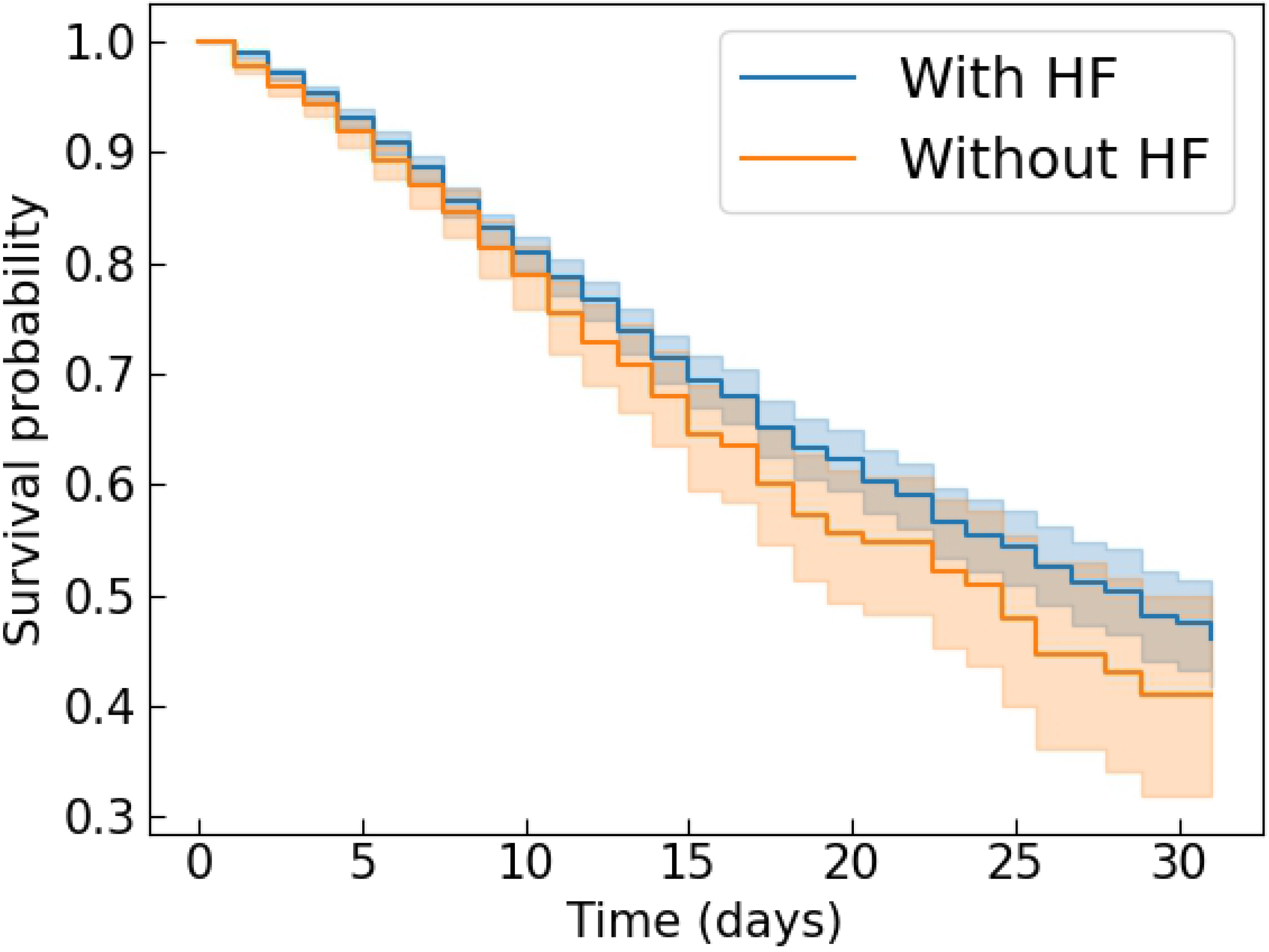
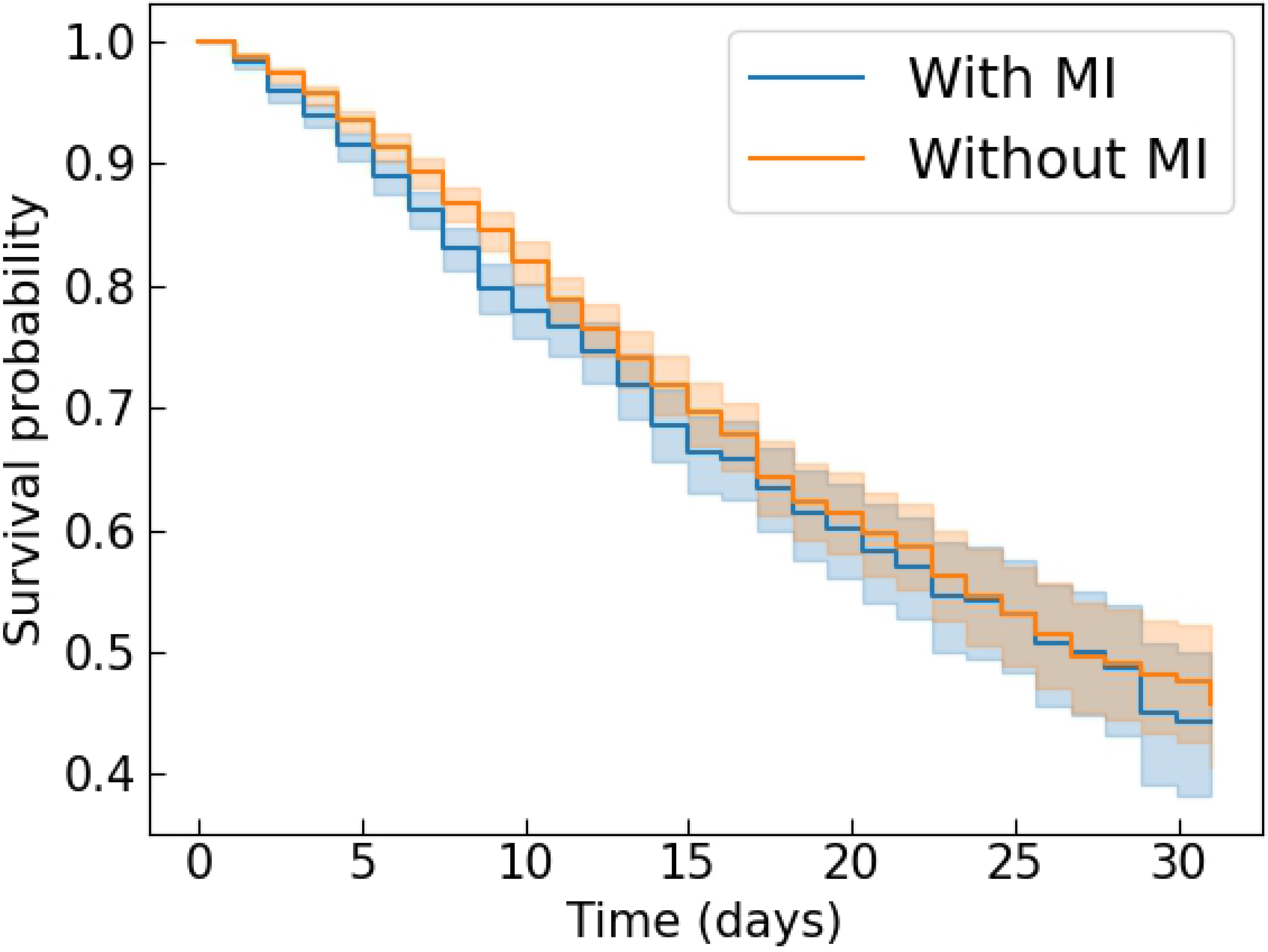
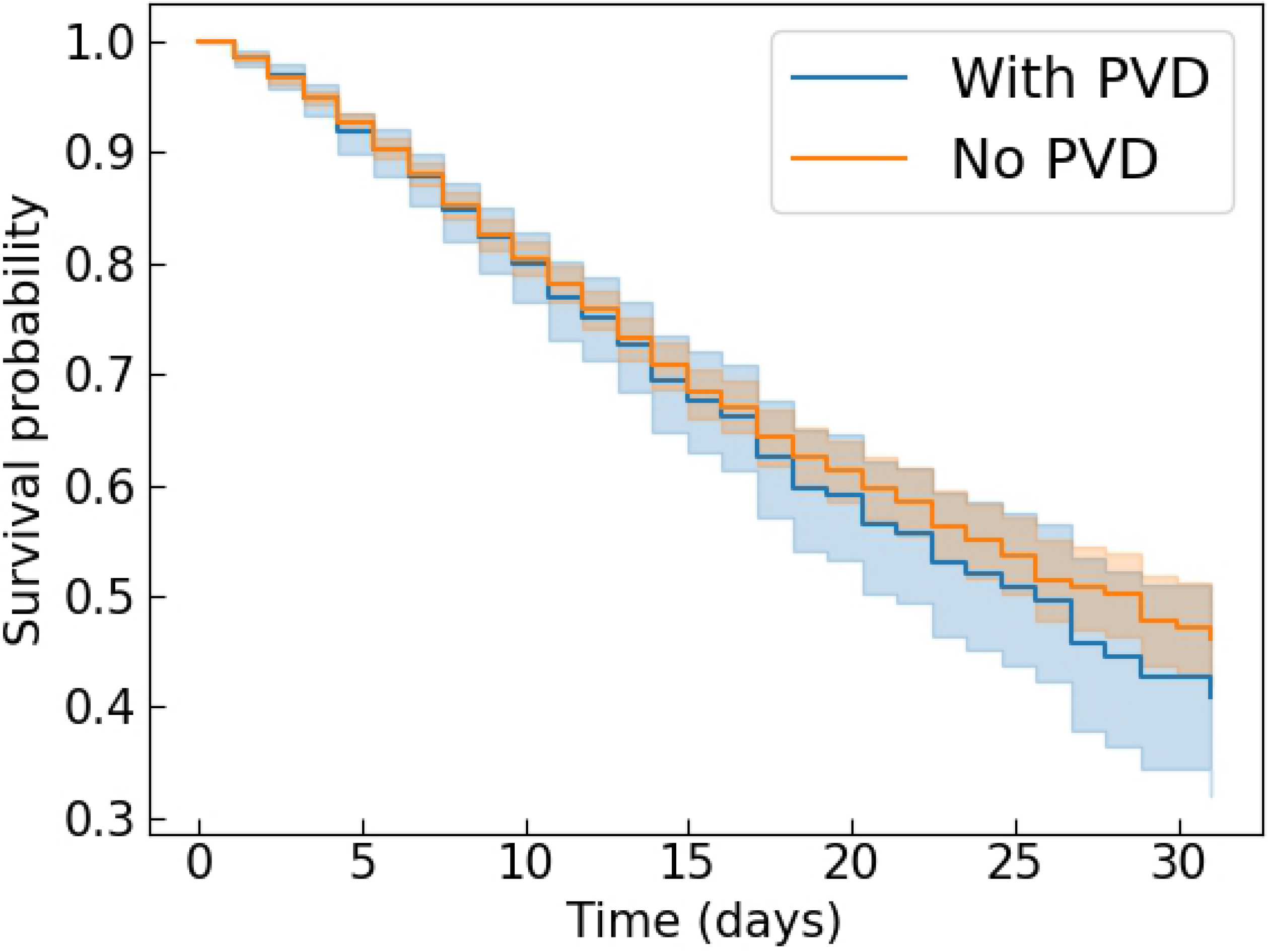
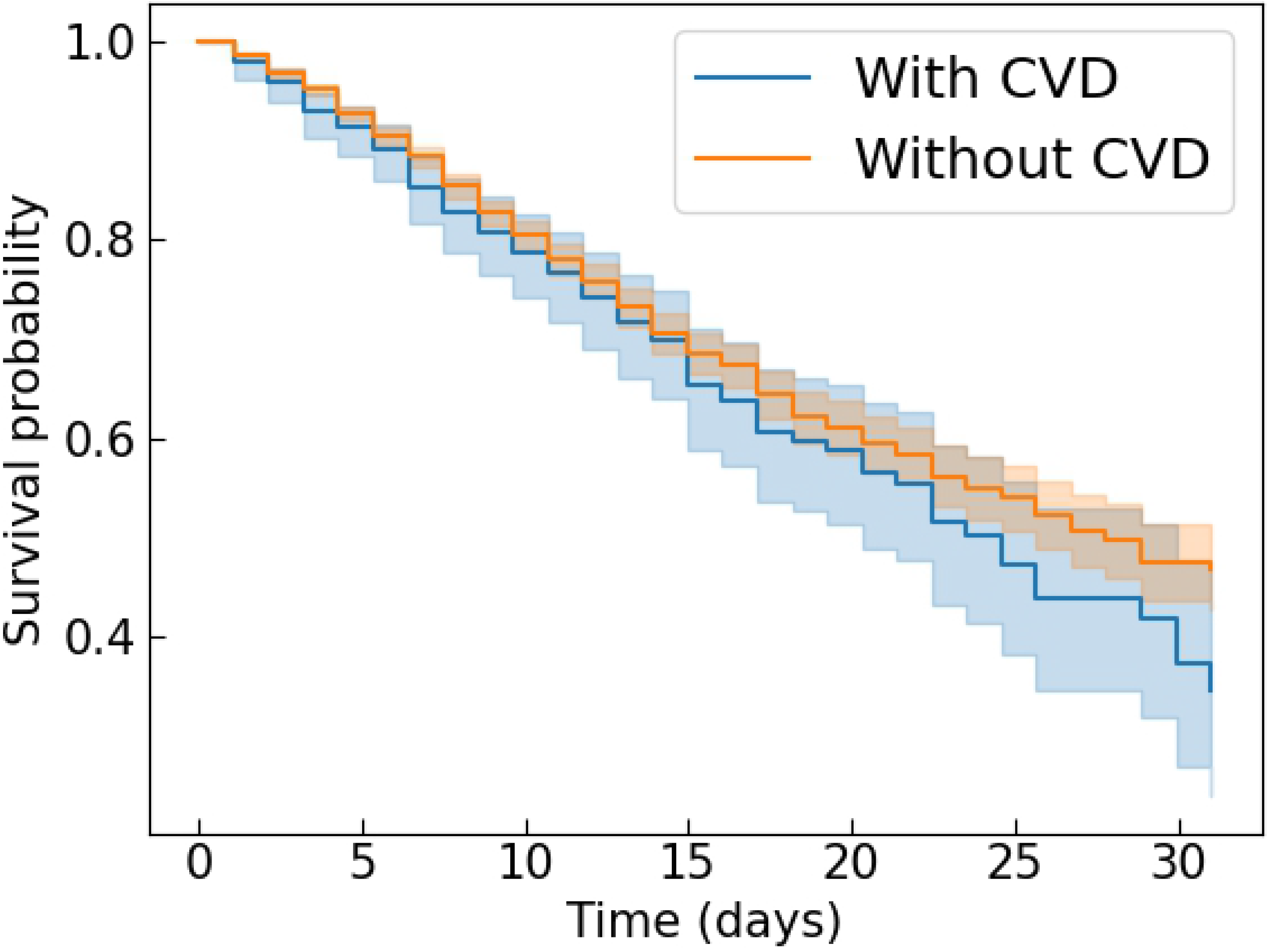
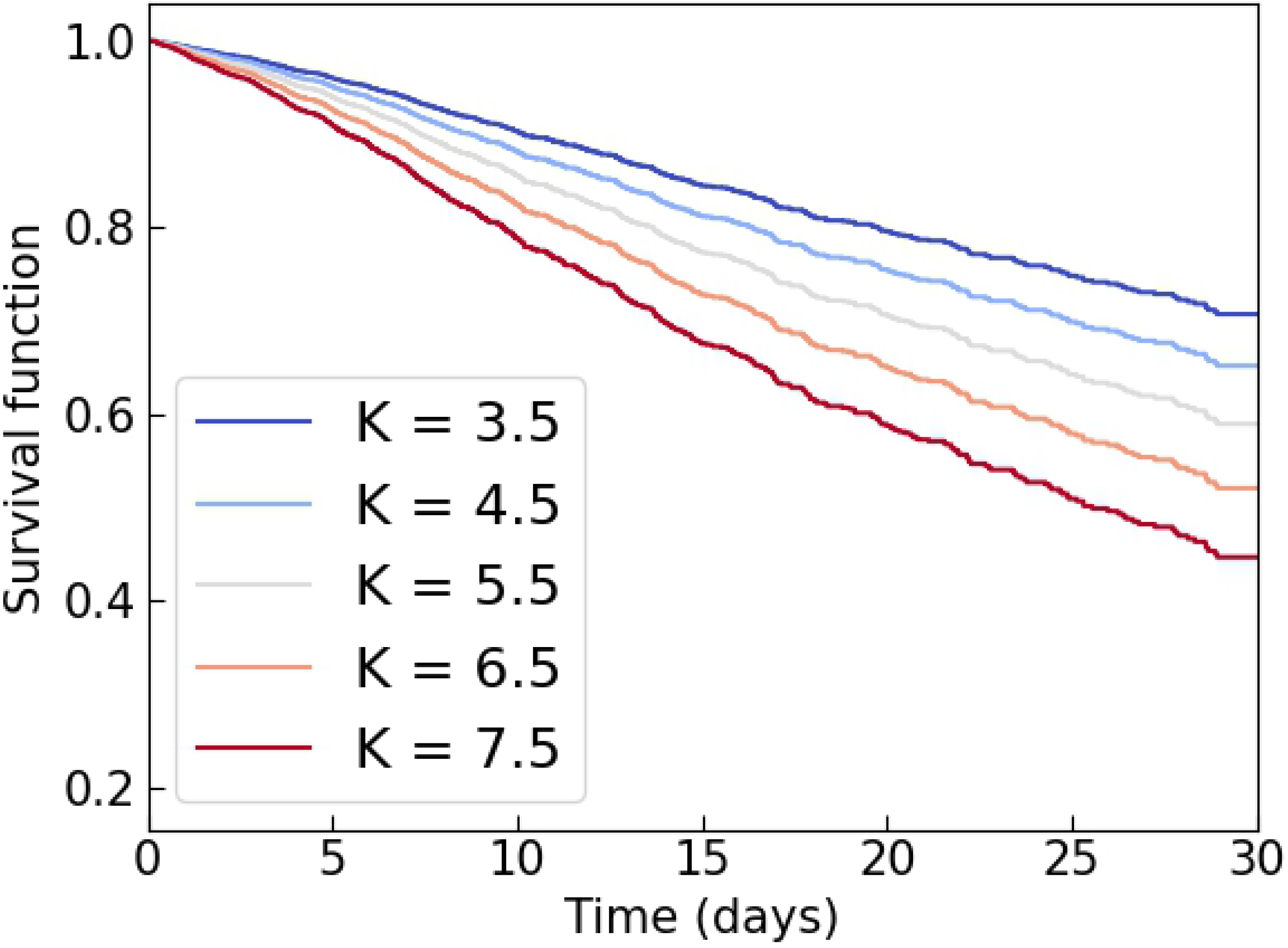
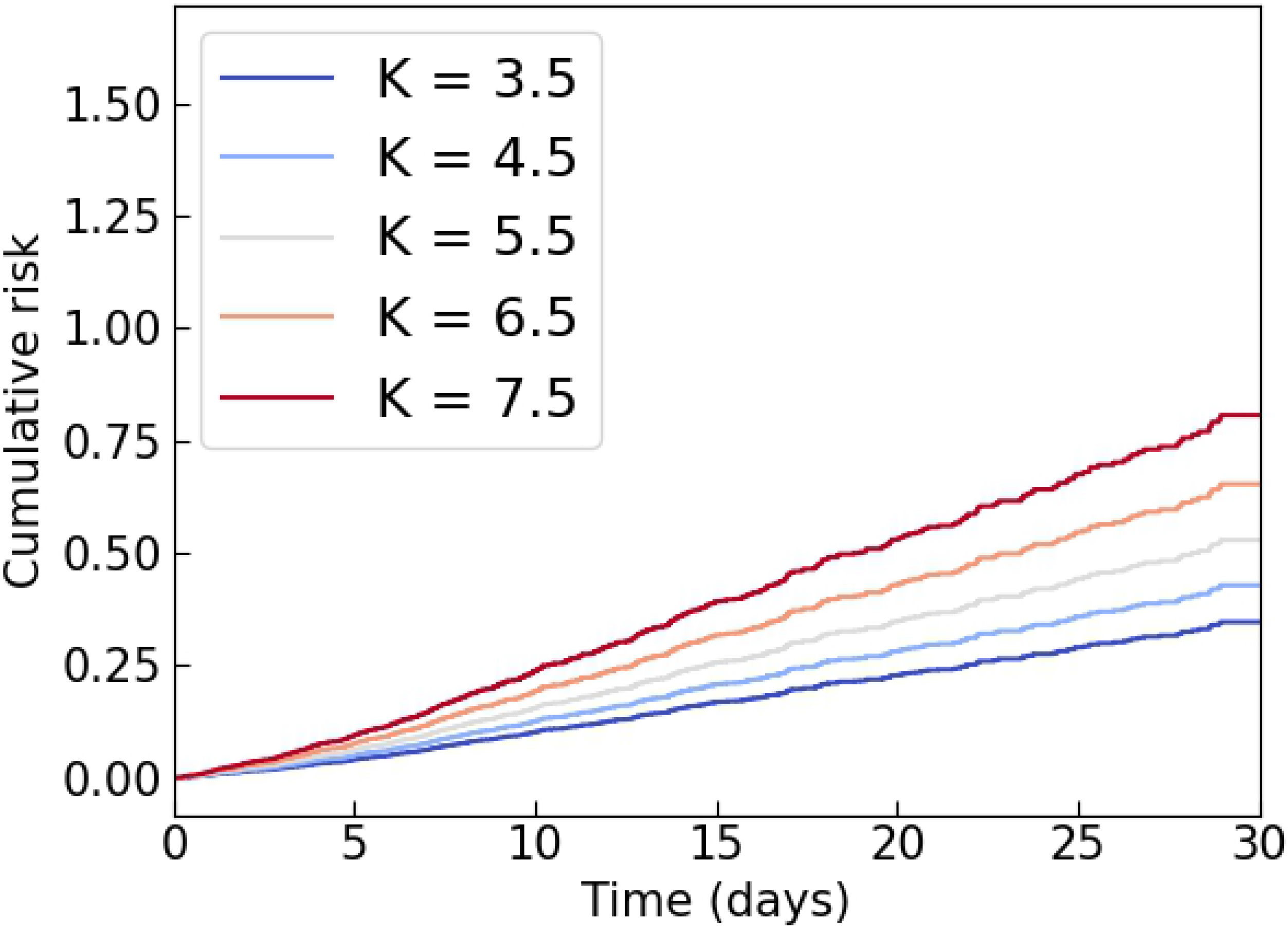
Survival and Cumulative Risk Functions allow us to observe that the higher the potassium level measured at intake is, compared to normal values, the lower the expected survival for the patient is.

This study suggested the association between increased levels of potassium, creatinine, hematocrit and white blood cells count with higher mortality in this CCU population.

In Cox’s model, by definition, no assumptions are made about the probability distribution of risk. In this case, the logarithmic risk of an individual was a linear function of its co-variables, plus a baseline risk of a given population that changes over time. For the purpose of this study, the adjusted Cox model was enough to demonstrate the importance of potassium levels in such patients in relation to their death.

Potassium disorders are prevalent in critically ill patients, and are also often found associated with cardiac comorbidities. In the case of hyperpotassemia, the causes may be multifactorial, and the main causes in this context are: (1) secondary to hormonal disorders, diabetes mellitus or chronic kidney disease; (2) drug-related (diuretics, renin-angiotensin-aldosterone system inhibitors, aldosterone receptor antagonists, non-steroidal anti-inflammatory or heparin); and (3) excessive potassium intake (whether dietary or supplemental). [15] In many cases, even though hyperpotassemia is a consequence or manifestation of another disorder, it will require medical intervention to avoid unfavorable outcomes. The findings of this article are in line with what was demonstrated by Brueske [16] in another cohort of CCU patients.

Creatinine was also related to shorter survival time in the population of this study, despite having a more discrete contribution than potassium. Previous studies have shown that small increases in creatinine are independently associated with higher mortality. [17–19]. This laboratory alteration is a key point in the current definitions of Acute Kidney Injury (AKI). [20] It has been demonstrated that AKI was independently associated with higher rates of incidence and progression of chronic kidney disease, as well as subsequent events of heart failure and death among survivors of recent hospitalization. [21]. The assessment of creatinine in this context is complex because it may represent a loss of previous renal function, concomitant and/or consequent to heart disease.

Increased hematocrit was associated with increased risk of death. This laboratory alteration is relatively frequent in the population of CCU and may be related to comorbidities (such as Chronic Obstructive Pulmonary Disease), a drug cause (hemoconcentration by the use of diuretics), and an inflammatory response. Blood cell disorders may influence higher risk of arterial and venous thrombosis, besides influencing the stability of existing thrombi. [22]

Leukocytosis is one of the most common laboratory changes in medicine and, in the patients studied, contributed with statistical significance to higher risk of death. It can have several causes, being the main infectious, inflammatory, and hematological disorders. [23] In relation to cardiovascular pathophysiology, the total increase in leukocytes is a risk factor for atherosclerosis, and is related to a higher incidence of coronary heart disease and ischemic stroke. Atherosclerosis has been shown to be a multifactorial disease with a basic inflammatory background. [24] Some clinical conditions of these patients, such as dehydration by diuretic use, can cause both leukocytosis and increased hematocrit.

Despite the aggregated value with the additional variables, although in this study population there was a statistically significant difference in NT-ProBNP levels between the groups of survivors and deaths, this variable was not a strong predictor by the regression model. In the case of Troponin T, the mean value between the two groups was equivalent and, likewise, there was no impact on mortality by the model presented. These two findings may be due to the fact that a significant portion of the patients did not present the measurement of this laboratory test in the first hours of hospitalization (only 15% and 50% for NT-ProBNP and Troponin T, respectively).

The results presented reinforced that the EHR contains relevant information to understand the progression of diseases and identify future directions for research. However, even when the distribution of a variable occurs according to a known physiological process, there may be a selective measurement, resulting in a non-physiological distributive representation of what should be a physiological variable. For this reason, the choice of the statistical model for the study of variables is a fundamental step and, when a simpler modeling technology is desired, Cox regression remains a good choice.

Despite the significant sample size and granular data, this is- everything has several limitations that are inherent to all retrospective cohort studies: missing data, a possible calibration bias, and the inability to adjust all possible relevant confounding factors. The loss of data may occur because the patient seeks care in another institution, or because of lack of standards in the filling of information.

This CCU population of a tertiary reference hospital may differ significantly from other populations. In this sense, the ICD-9 discharge codes that we obtained reflect all hospitalization diagnosis, including acute and chronic conditions.

The EHR data are complex and present several points of attention: specificities of the cohort of patients and its impact on the generalization of the analyses, absent data in a non-random way, large number of dimensions to represent patient information with unknown interrelationships that make the process of variable selection complex, and data collection uncontrolled. All these factors may interfere with the results of the proposed models.

There was also an additional limitation in this study, which was the unavailability of MIMIC-IV clinical notes. For this reason, relevant information such as metrics obtained by the Transthoracic Ecodooplercardiogram could not be included in the study.

The importance of conducting cohort studies with patients hospitalized in CCU with longer follow-up time, including post-discharge, could be relevant to evaluate the quality and years of life aggregated after the treatment offered to patients.

## Conclusion

The study of this cohort of CCU patients has allowed us to suggest associations between clinical variables (vital signs on admission and frequent comorbidities) and higher risk of in-hospital death. The results have indicated that survival analysis combined with EHR data may be possible when data are treated appropriately, and may highlight factors, both clinical and related to health indicators, who pose a higher risk of death.

The main contributions of this article were: (i) clear data processing, in order to allow the reproduction of this and other research in the MIMIC-IV database and mitigate doubts regarding concepts, missing data and censorship; (ii) the comparison of two survival regression models applicable in EHR data; and (iii) the identification of the importance of monitoring potassium levels in this group of patients for early identification of indications of unfavorable outcome.

In this group of patients, elderly people represent a significant number of hospitalizations in CCU. This may be associated with a different profile of comorbidities that represent a higher risk of death and can be explored in future studies. The survival analysis of other groups of MIMIC-IV patients is also a research opportunity that has not yet been explored. New studies may also determine ideal models with a larger set of variables, including laboratory tests, comorbidities, medications, and other potentially relevant characteristics of the patient.

The results of this article, along with additional research on the generalization of known risk indicators among critical ICU patients, such as APACHE [25], SOFA [26], LODS [27], OASIS [28] and SAPS II [29], may provide evidence to guide the use of more appropriate risk scores for the population of CCUs.

## Data Availability

The data underlying the results presented in the study are available from Physionet. It can become available from the MIT Institutional Data Access (contact via https://physionet.org/about/) for researchers who meet the criteria for access to confidential data.

## Supporting information

## Acknowledgments

We would like to thank the editors and reviewers of PLoS ONE for their thoughtful comments.

## References

1. Kasaoka S. Evolved role of the cardiovascular intensive care unit (CICU). Journal of Intensive Care. 2017;5(1). doi:10.1186/s40560-017-0271-7.

2. Dai Z, Liu S, Wu J, Li M, Liu J, Li K. Analysis of adult disease characteristics and mortality on MIMIC-III. PLOS ONE. 2020;15(4):e0232176. doi:10.1371/journal.pone.0232176.

3. Kadooka K, Miyachi H, Kimura T, Asano K, Onodera K, Masunaga N, et al. Non-cardiovascular disorders in a contemporary cardiovascular intensive care unit in Japan. Journal of Cardiology. 2021;78(2):166–171. doi:10.1016/j.jjcc.2021.03.002.

4. Jentzer JC, Anavekar NS, Bennett C, Murphree DH, Keegan MT, Wiley B, et al. Derivation and Validation of a Novel Cardiac Intensive Care Unit Admission Risk Score for Mortality. Journal of the American Heart Association. 2019;8(17). doi:10.1161/jaha.119.013675.

5. Bohula EA, Katz JN, van Diepen S, Alviar CL, Baird-Zars VM, Park JG, et al. Demographics, Care Patterns, and Outcomes of Patients Admitted to Cardiac Intensive Care Units. JAMA Cardiology. 2019;4(9):928. doi:10.1001/jamacardio.2019.2467.

6. Lighthall GK, Vazquez-Guillamet C. Understanding Decision Making in Critical Care. Clinical medicine & research. 2015;13(3-4):156–168. doi:10.3121/cmr.2015.1289.

7. Kane RL, Shamliyan TA, Mueller C, Duval S, Wilt TJ. The Association of Registered Nurse Staffing Levels and Patient Outcomes. Medical Care. 2007;45(12):1195–1204. doi:10.1097/mlr.0b013e3181468ca3.

8. Baker S, Xiang W, Atkinson I. Continuous and automatic mortality risk prediction using vital signs in the intensive care unit: a hybrid neural network approach. Scientific Reports. 2020;10(1). doi:10.1038/s41598-020-78184-7.

9. Rosenberg AL, Watts C. Patients Readmitted to ICUs: A Systematic Review of Risk Factors and Outcomes. Chest. 2000;118(2):492–502. doi:https://doi.org/10.1378/chest.118.2.492.

10. Singh R, Mukhopadhyay K. Survival analysis in clinical trials: Basics and must know areas. Perspectives in clinical research. 2011;2(4):145–148. doi:10.4103/2229-3485.86872.

11. Harerimana G, Kim JW, Jang B. A deep attention model to forecast the Length Of Stay and the in-hospital mortality right on admission from ICD codes and demographic data. Journal of Biomedical Informatics. 2021;118:103778. doi:https://doi.org/10.1016/j.jbi.2021.103778.

12. Johnson A, Bulgarelli L, Pollard T, Horng S, Celi LA, Mark R. MIMIC-IV; 2020. Available from: https://physionet.org/content/mimiciv/0.4/.

13. Davidson-Pilon C. lifelines: survival analysis in Python. Journal of Open Source Software. 2019;4(40):1317.

14. Pollard TJ, Johnson AEW, Raffa JD, Mark RG. tableone: An open source Python package for producing summary statistics for research papers. JAMIA Open. 2018;1(1):26–31. doi:10.1093/jamiaopen/ooy012.

15. Sarwar CMS, Papadimitriou L, Pitt B, Pinã I, Zannad F, Anker SD, et al. Hyperkalemia in Heart Failure. Journal of the American College of Cardiology. 2016;68(14):1575–1589. doi:https://doi.org/10.1016/j.jacc.2016.06.060.

16. Brueske B, Sidhu MS, Schulman-Marcus J, Kashani KB, Barsness GW, Jentzer JC. Hyperkalemia Is Associated With Increased Mortality Among Unselected Cardiac Intensive Care Unit Patients. Journal of the American Heart Association. 2019;8(7). doi:10.1161/jaha.118.011814.

17. Thomas ME, Blaine C, Dawnay A, Devonald MAJ, Ftouh S, Laing C, et al. The definition of acute kidney injury and its use in practice. Kidney International. 2015;87(1):62–73. doi:10.1038/ki.2014.328.

18. Lassnigg A. Minimal Changes of Serum Creatinine Predict Prognosis in Patients after Cardiothoracic Surgery: A Prospective Cohort Study. Journal of the American Society of Nephrology. 2004;15(6):1597–1605. doi:10.1097/01.asn.0000130340.93930.dd.

19. Lassnigg A, Schmid ER, Hiesmayr M, Falk C, Druml W, Bauer P, et al. Impact of minimal increases in serum creatinine on outcome in patients after cardiothoracic surgery: Do we have to revise current definitions of acute renal failure? Critical Care Medicine. 2008;36(4):1129–1137. doi:10.1097/ccm.0b013e318169181a.

20. Khwaja A. KDIGO Clinical Practice Guidelines for Acute Kidney Injury. Nephron. 2012;120(4):c179–c184. doi:10.1159/000339789.

21. Ikizler TA, Parikh CR, Himmelfarb J, Chinchilli VM, Liu KD, Coca SG, et al. A prospective cohort study of acute kidney injury and kidney outcomes, cardiovascular events, and death. Kidney International. 2021;99(2):456–465. doi:10.1016/j.kint.2020.06.032.

22. Byrnes JR, Wolberg AS. Red blood cells in thrombosis. Blood. 2017;130(16):1795–1799. doi:10.1182/blood-2017-03-745349.

23. Cerny J, Rosmarin AG. Why Does My Patient Have Leukocytosis? Hematology/Oncology Clinics of North America. 2012;26(2):303–319. doi:10.1016/j.hoc.2012.01.001.

24. Haybar H, Pezeshki SMS, Saki N. Evaluation of complete blood count parameters in cardiovascular diseases: An early indicator of prognosis? Experimental and Molecular Pathology. 2019;110:104267. doi:10.1016/j.yexmp.2019.104267.

25. Knaus WA, Wagner DP, Draper EA, Zimmerman JE, Bergner M, Bastos PG, et al. The APACHE III Prognostic System. Chest. 1991;100(6):1619–1636. doi:10.1378/chest.100.6.1619.

26. Vincent JL, Moreno R, Takala J, Willatts S, De Mendonca A, Bruining H, et al. The SOFA (Sepsis-related Organ Failure Assessment) score to describe organ dysfunction/failure. On behalf of the Working Group on Sepsis-Related Problems of the European Society of Intensive Care Medicine. Intensive care medicine. 1996;22(7):707–10.

27. Gall JRL. The Logistic Organ Dysfunction System. JAMA. 1996;276(10):802. doi:10.1001/jama.1996.03540100046027.

28. Johnson AEW, Kramer AA, Clifford GD. A New Severity of Illness Scale Using a Subset of Acute Physiology and Chronic Health Evaluation Data Elements Shows Comparable Predictive Accuracy. Critical Care Medicine. 2013;41(7):1711–1718. doi:10.1097/ccm.0b013e31828a24fe.

29. Gall JRL. A new Simplified Acute Physiology Score (SAPS II) based on a European/North American multicenter study. JAMA: The Journal of the American Medical Association. 1993;270(24):2957–2963. doi:10.1001/jama.270.24.2957.

